# Modelling practices, data provisioning, sharing and dissemination needs for pandemic decision-making: a European survey-based modellers’ perspective

**DOI:** 10.1101/2025.03.12.25323819

**Authors:** Esther van Kleef, Wim Van Bortel, Elena Arsevska, Luca Busani, Simon Dellicour, Laura Di Domenico, Marius Gilbert, Sabine L. van Elsland, Moritz U.G. Kraemer, Shengjie Lai, Philippe Lemey, Stefano Merler, Zoran Milosavljevic, Annapaola Rizzoli, Danijela Simic, Andrew J. Tatem, Maguelonne Teisseire, William Wint, Vittoria Colizza, Chiara Poletto

## Abstract

**Introduction:** Advanced outbreak analytics played a key role in governmental decision-making as the COVID-19 pandemic challenged health systems globally. This study assessed the evolution of European modelling practices, data usage, gaps, and interactions between modellers and decision-makers to inform future investments in epidemic-intelligence globally.

**Methods:** We conducted a two-stage semi-quantitative survey among modellers in a large European epidemic-intelligence consortium. Responses were analysed descriptively across early, mid-, and late-pandemic phases. Policy citations in Overton were used to assess the policy impact of modelling.

**Findings:** Our sample included 66 modelling contributions from 11 institutions in four European countries. COVID-19 modeling initially prioritised understanding epidemic dynamics, while evaluating non-pharmaceutical interventions and vaccination impacts became equally important in later phases. ‘Traditional’ surveillance data (e.g. case linelists) were widely used in near-real time, while real-time non-traditional data (notably social contact and behavioural surveys), and serological data were frequently reported as lacking. Data limitations included insufficient stratification and geographical coverage. Interactions with decision-makers were commonplace and informed modelling scope and, vice versa, supported recommendations. Conversely, fewer than half of the studies shared open-access code.

**Interpretation:** We highlight the evolving use and needs of modelling during public health crises. The reported missing of non-traditional surveillance data, even two years into the pandemic, underscores the need to rethink sustainable data collection and sharing practices, including from for-profit providers. Future preparedness should focus on strengthening collaborative platforms, research consortia and modelling networks to foster data and code sharing and effective collaboration between academia, decision-makers, and data providers.

## Introduction

The COVID-19 pandemic represented one of the most significant global health emergencies in recent years.^1^ While the immediate threat of COVID-19 has receded, the widespread circulation of Mpox clades I and II, alongside the escalating risk from influenza H5N1, underscores the continuous threat of viral emergence events and the importance of investment in preparedness for new pandemic threats.^2^

During the COVID-19 crisis, governments and policymakers implemented detection, monitoring, and intervention strategies in the face of an uncertain and rapidly evolving epidemiological landscape.^3^ As such, COVID-19 exposed vulnerabilities in public health systems worldwide. To support decision-making, mathematical and computational approaches - commonly referred to as ‘outbreak analytics’ - played a critical role.^4–6^ This was paralleled by an unprecedented effort in the collection and sharing of both epidemiological data such as case reporting, viral genome sequencing and serological data, and ‘non-traditional’ surveillance data, e.g. mobility flows reconstructed by mobile phones, geolocation and air-travel data, social-mixing surveys, and sentiment data from social media^7^.

Reflections and commentaries by scientists, alongside high-level multilateral and expert reviews, have provided key insights into modelling-specific lessons learned.^7–9^ The discourse has focused on identifying modelling and data needs at different outbreak stages,^6,7,10–20^ developing efficient and flexible data collection frameworks that can rapidly scale up when necessary,^21,22^ improving communication and collaboration between modellers and public health authorities,^9,23–27^ and a rethinking of rewarding structures and institutional support for science-policy activities.^28^

Here, we contribute to these reflections with an objective and systematic reconstruction of the outbreak analytics activities conducted throughout the COVID-19 pandemic by MOOD (MOnitoring Outbreaks for Disease Surveillance in a data science context) - a large, multi-partner, multi-country epidemic intelligence consortium (mood-h2020.eu). We identified how outbreak analytics’ scope, methodologies, and input data evolved throughout the pandemic. We examined data sources, data limitations, and missing data. We then reviewed the nature of interactions between scientists and decision-makers and the policy impact of modelling. Hence, this study provides insights into how quantitative scientific advice evolved, the extent to which it was implemented in policy, and the gaps in perceived needs between scientists and policymakers.

## Methods

### Study participants

The MOOD project is a Horizon2020 EU-funded project (start date January 2020, end date December 2024), bringing together partners from academic, research, public health, and animal health institutions. The project aimed to develop innovative tools and services for the early detection, assessment, and monitoring of current and potential infectious disease threats in EU/EEA using a participatory approach^29^. Modelling teams within MOOD were located in four countries with different socio-ecological (Nordic, Western, Central, and Mediterranean environments) and epidemiological (organisation and structure of health services, state of health infrastructures, complexity of surveillance systems) characteristics. Hence, partners of the MOOD consortium and the application of their work provided a broad representation of the European region.

### Survey

Following a two-stage approach, we developed a semi-quantitative survey among MOOD partners. In April 2022, we conducted a scoping survey based on open-ended questions. We inventoried all modelling studies on the COVID-19 pandemic until the survey date, here defined as encompassing computational (e.g., machine learning and phylogenetic approaches), statistical (e.g., spatial-temporal models, regression techniques) and mechanistic modelling (e.g., compartmental and agent-based models) studies. Based on answers in the scoping survey, we designed a final semi-quantitative survey using similar inclusion criteria and predominantly closed questions (full survey in the supplementary material). The survey was distributed among MOOD partners in July 2023. All studies carried out before the survey date were eligible for inclusion. These included studies conducted in collaboration with or assigned by public health authorities (PHA) and/or decision-makers, as well as those communicated to PHA (either directly or indirectly, e.g., via scientific publication) or neither.

The final survey covered questions on: (1) Objectives of the implemented work, (2) Methodologies used; (3) Data most frequently used and missing; (4) Data availability and access and reason for lack of access; (5) Means of collaboration between scientists and decision-makers. In addition, for each study, the survey inventoried the pandemic period to which the study was referring (year and month of beginning and year and month of end), code-sharing practices, the geographical scope of the work, and the generalisability of the study to other diseases or geographical locations. Participants could select answers from a predefined list, with answers not mutually exclusive in many cases and the possibility to add free text to specify answers not included.

### Analyses

Free text answers were manually analysed, and response categories were re-coded where relevant. We classified studies according to whether their end date was aligned with the early, mid- or later pandemic phases, defined as 1) between January and June 2020 (dominating variant: wild type), 2) between July 2020 and June 2021 (Alpha wave), and 3) after July 2021 (Delta and Omicron waves), respectively. Then, analyses involved descriptive statistics, cross-tabulation, and stratification by pandemic period. We applied Fisher’s exact test to assess differences in modelling scope and data completeness by the degree and nature of science-policy interactions. As differences were tested for each modeling scope and data type separately, we applied a Bonferroni correction to adjust for multiple comparisons and control the type I error rate.

Code used for the analyses and figures is available on GitHub (Data sharing statement below). To measure policy impact, we used as a metric the number of policy documents that cited the respective modelling works identified in the Overton database^5^. Overton is a global database of national and intergovernmental policy documents that allows for tracking of policy citations, including citations to scientific evidence of interest. All Overton reports were extracted on 19-12-2024.

## Ethical approval

The Institutional Review Board of the Institute of Tropical Medicine Antwerp approved the study protocol as part of an overarching qualitative study (ref no. 1484/21). All data were collected by MOOD project partners with their consent upon filling out the survey.

## Results

The final survey resulted in a sample of 66 modelling contributions by 11 institutions from four European countries (France, Belgium, Italy, and the United Kingdom). Missing records amount to 3%. After manually resolving ambiguities, we assigned a pandemic phase to each of the contributions. Our sample included a close to equal number of studies for each pandemic phase, i.e. 24, 21 and 21 studies from the early-, mid-, and later-pandemic phases, respectively. The geographical scope was defined for 65 out of 66 studies and notably included national (n=24) and sub-national level (n=19, Figure 1), followed by global (n=16) and continent-level (n=5). The modelling work was considered applicable to other geographical locations for all works but two - in different locations with similar socio-demographic contexts for around one-third of studies (n=21) and everywhere worldwide for two-thirds (n=43). Finally, most works were considered applicable to other diseases (63/66, 95%). Around a third of the works (32%) leveraged openly available code. Conversely, 47% of the studies (n=31) developed code that was made publicly available, with GitHub as a predominant outlet.

**Figure 1.**
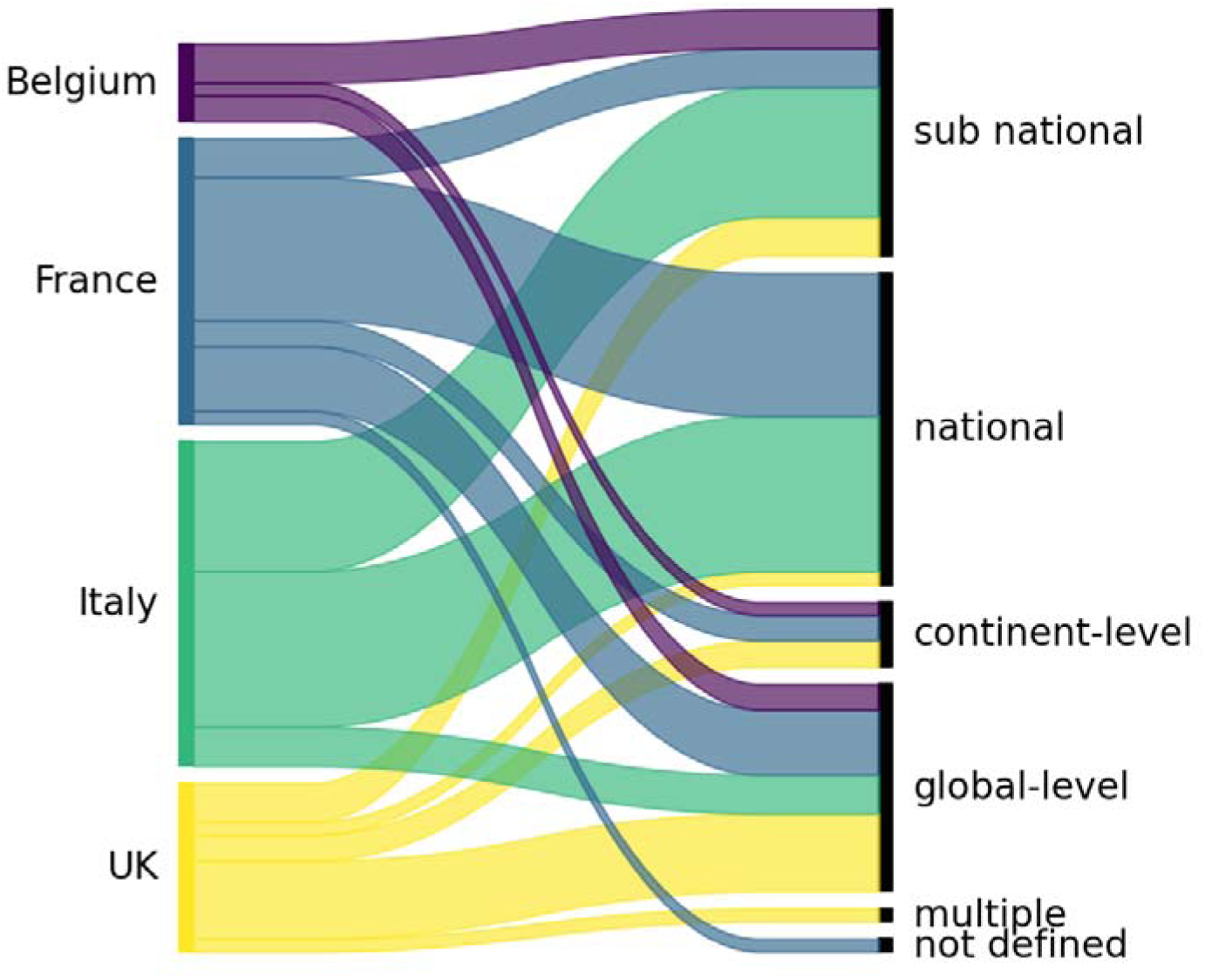
Geographical scope of the work. Alluvial diagram of the number of surveyed works by country vs. geographical scope.

### Scope and analytical methods in the early and later COVID-19 pandemic phases (January 2020 to post-July 2021)

Overall, modelling work during the COVID-19 pandemic provided an understanding of COVID-19 dynamics (n=43, Figure 2A), that is, the estimation of transmission parameters (n=14), quantification of the COVID-19 burden (n=12), understanding of determinants of geographical spread (n=7), estimation of the true number of cases/under-reporting (n=8), and others (e.g., clinical aspects, emergence). Secondly, modelling assessed the impact of non-pharmaceutical interventions (NPIs, n=34), including lockdown (n=15), social distancing (n=15), and travel restrictions (n=14). Understanding the epidemic dynamics remained the most important, or among the most important, focus throughout the pandemic. In addition, the fraction of studies evaluating the impact of NPIs increased in the mid-pandemic phase, while the evaluation of vaccination became notable in the later-pandemic phase (after June 2021).

**Figure 2.**
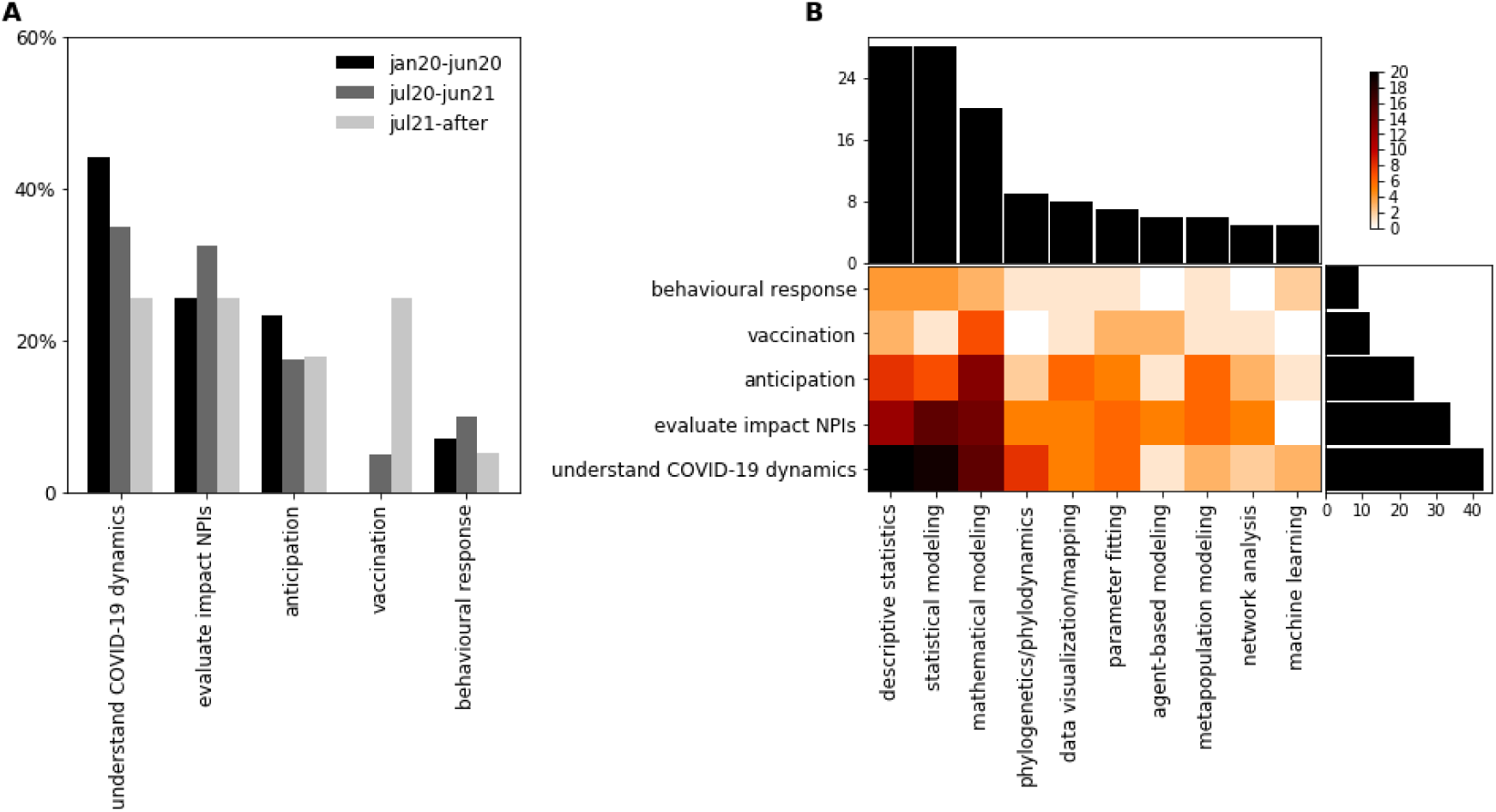
Scope and methodology of modelling works. **A** Frequency of the different goals across the three phases of the pandemic. **B** number of different goals and adopted methodologies.

Descriptive statistics and statistical modelling were frequently employed (n=28, both cases), primarily to understand the COVID-19 transmission dynamics (Figure 2B). This was followed by mathematical modelling (n=20), which was the preferred approach for studying the impact of vaccination and anticipating the course of the pandemic (Figure 2B).

### Data (re)sources

The most widely adopted epidemiological data types included case line lists (n=32), e.g. to estimate epidemiological transmission parameters such as generation time, basic-/ net-reproduction number, and incubation time (n=9), and quantify the burden of COVID-19 (n=8), followed by COVID-19 incidence data (n=24, and genomic data, (n=15, Figure 3A). Furthermore, mobility, n=56 (grouping all possible types of mobility data), socio-economic data, n=33 (e.g. socio-economic indicators, social contacts, etc.), and population characteristics, n=20 (including age, gender, ethnicity, comorbidities) were frequently sourced. Pre-pandemic and real-time mobility were used to inform models, notably during the early- and mid-pandemic phases (Figure 3A). Conversely, vaccination, attitude/behavioural surveys, and social structure (household structure, school catchments, workplace size, and distribution) were specifically sourced and used in the mid- and later phases.

**Figure 3.**
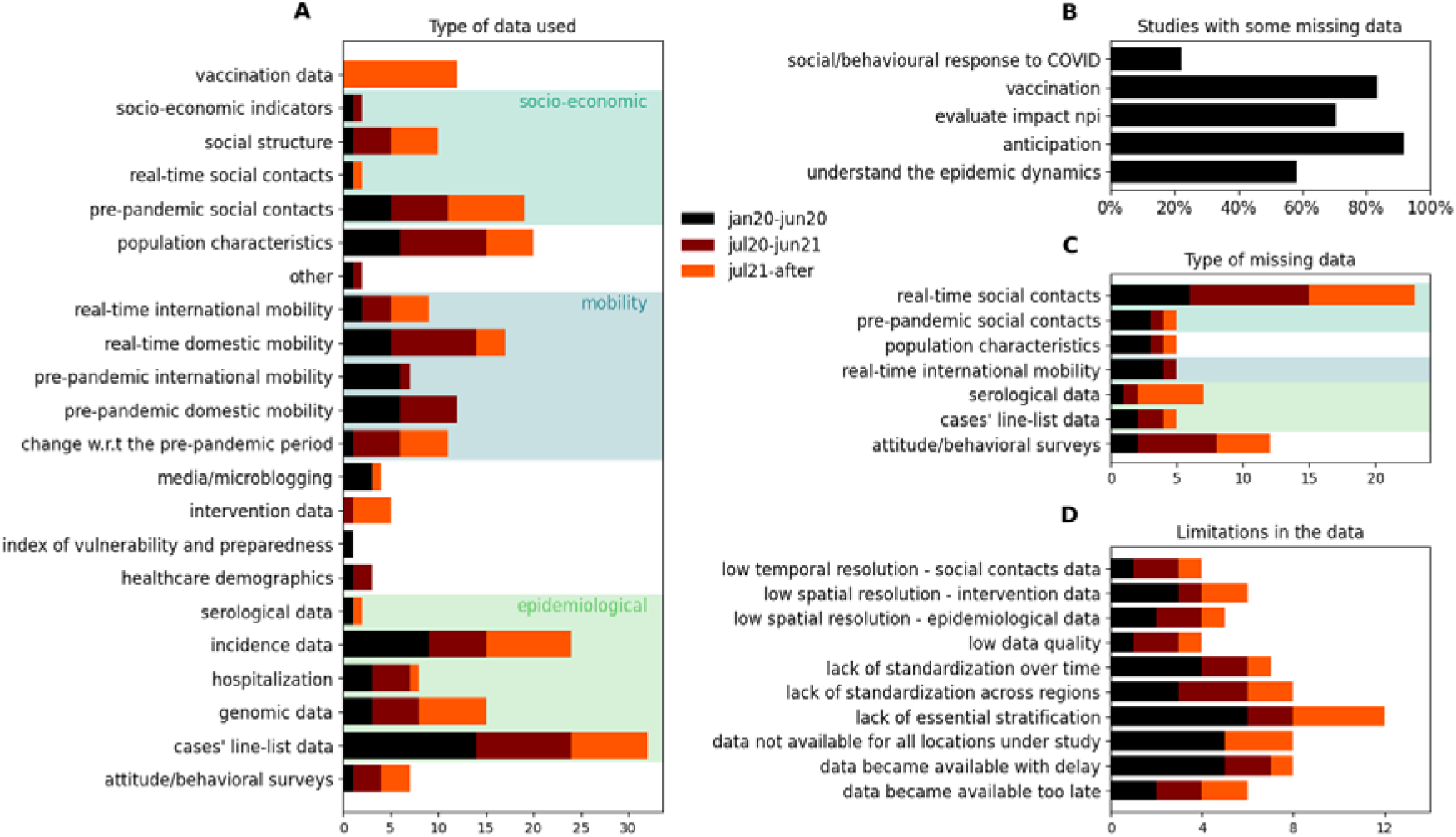
Data types used vs missing. **A** Stacked histogram of data types used stratified by pandemic phases - statistics over 65 works, one study used no data. Epidemiological, mobility and socio-economic data are grouped. **B** Proportion of studies with missing data for each study goal. **C** Stacked histogram of missing data types stratified by pandemic phase. Only data with an aggregated count of five or more are displayed. Epidemiological, mobility and socio-economic data are grouped with the same colour code as in panel A. **D** Stacked histogram of data limitations stratified by pandemic phase. Only data with an aggregated count of four or more are displayed.

Overall, the data used became available thanks to the collecting and sharing in near real-time by PHA (n=45), were gathered before the pandemic from freely and openly available sources (n=25), or were collected specifically for and in the context of the modelling work of concern by study collaborators (n=20). Interestingly, for-profit organisations were relevant data providers during the COVID-19 pandemic, sharing their data directly with modellers for a quarter of the studies (n=16) or making the data openly available in the context of data-for-good initiatives (n=16). This was the same for data shared through academic initiatives or initiatives of non-profit organisations (n=15).

### Data missing and access

For over half of the modelling works, missing relevant data was reported (n=40/66, 61%). This percentage slightly increased throughout the different phases of the pandemic, i.e. from 54% (n=13/24) in the early phase to 67% (n=14/21) in the later pandemic phase. Missing data affected primarily studies aimed at anticipating the course of the pandemic (n=22/24, 92%), followed by those assessing the impact of vaccination (n=10/12, 83%) or NPIs (n=24/34, 71%) (Figure 3B). Particularly lacking were real-time social contact data (n=23) and data on attitudes and behaviours (n=12, Figure 3C). The reported relative lack of these data remained similar throughout the pandemic.

The main reasons listed for missing data were that the data was never collected (n=27), the process for obtaining them was too lengthy (n=9), or the data were protected by privacy or ethical restrictions (n=7). Where data were available, limitations were faced in 39 studies, most notably related to a lack of essential stratification (e.g. incidence by age, sex, and comorbidity, n=12), delay in data availability (n=8), lack of available data for all locations under study (n=8), or lack of standardisation across regions (n=8, Figure 3D).

### Interaction with Public Health Authorities

For most modelling contributions (n=58/66, 88%), survey participants listed that their work supported the scientific understanding or situational awareness of decision-makers or PHA (Figure 4A). For a smaller but marked fraction, participants perceived that their work supported PHA official recommendations (n=51/66, 77%) and/or was directly prompted by a discussion with PHA (n=41/66, 62%). Direct interactions with PHAs occurred for 54 out of 66 studies and involved direct collaboration (n=45/54, 82%), discussions at internal meetings (n=4/54, 7%), or participation of modellers in advisory committees (n=5/54, 9%). Direct collaborations were noted particularly during the early-pandemic phase (75%, 66%, and 62% reporting direct collaboration with PHA in the early-, mid-, and later-pandemic phases respectively), while interactions through advisory committees occurred during the mid- and later pandemic phases (0%, 14%, and 10% reporting interaction through advisory committees in the early-, mid-, and later-pandemic phases respectively).

**Figure 4.**
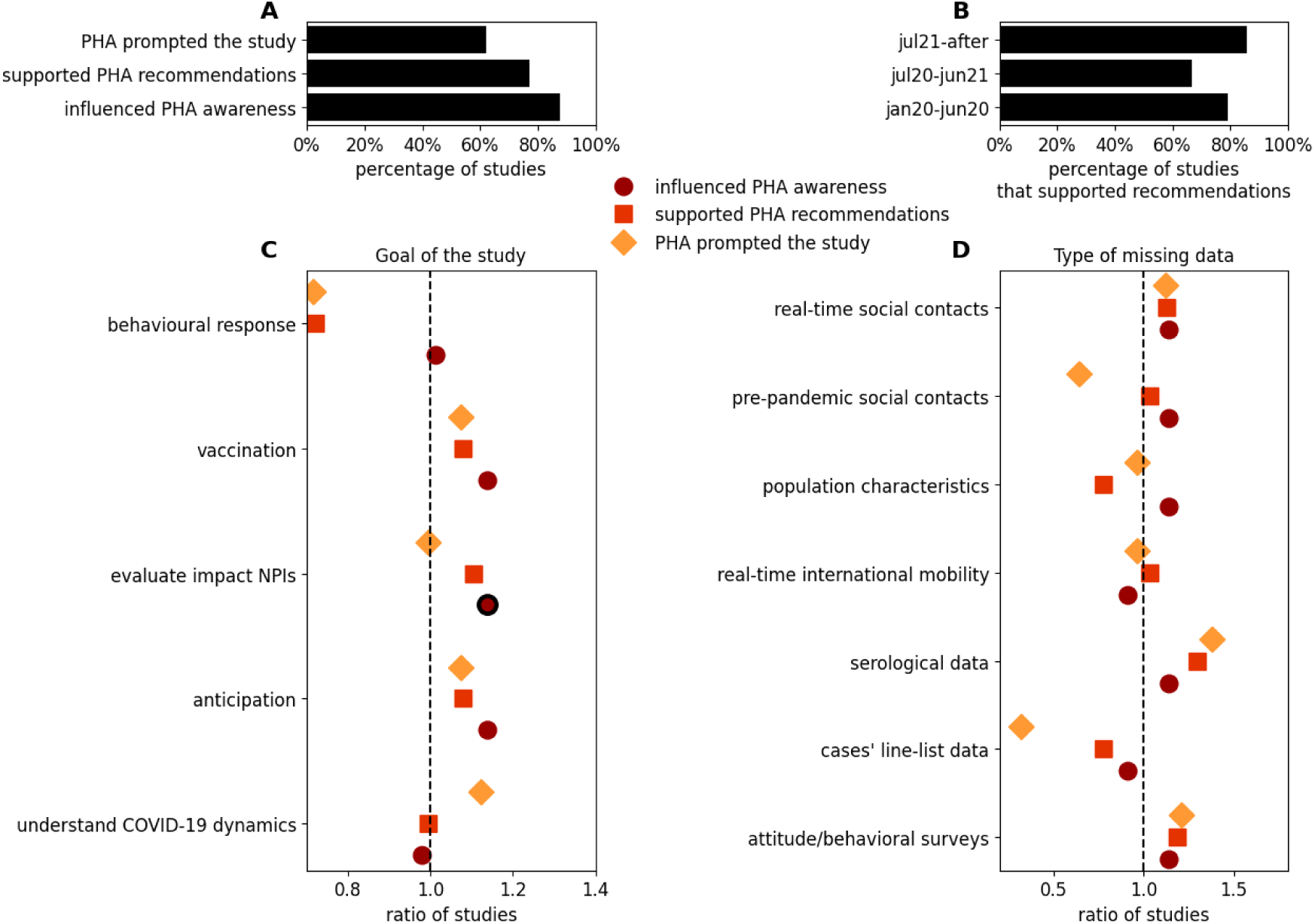
Interaction with Public Health Authorities (PHA). **A** Proportion of studies with a certain type of outreach toward PHAs. We consider three kinds of outreach: 1) the study influenced the scientific understanding or situational awareness of PHA, 2) supported official COVID-19 recommendations by PHA, 3) was prompted by discussions with PHA. **B** Percentage of studies that supported official COVID-19 recommendations by PHA in each pandemic phase. **C** Ratio of the proportion of studies with a particular scope among each of three different possible interactions with PHA divided by the proportion of studies with that same objective across all studies (e.g. 73% of the studies prompted by PHA concerned understanding the epidemic dynamics, while this was 65% across all studies, which results in a ratio of 1.12). Black thick border indicated statistical significance based on Fisher’s exact test, with p<0.05 after correction for multiple testing. **D:** same statistic as in C, but for types of missing data.

Among the 58 studies that supported situational awareness of PHA, there was a higher percentage of studies that evaluated the impact of NPIs (n=34/58, 59%), compared to their relative representation among all the modelling studies (i.e. including the studies that did not report an interaction with PHA, n= 34/66, 52%, Fisher’s exact test p<0.05, after correcting for multiple comparisons, Figure 4C).

The number of studies supporting official recommendations was the highest in the later-pandemic phase immediately followed by the early one (Figure 4B). Analysing the relative representation of missing data across studies according to their outreach toward PHAs (Figure 4D) illustrated that the lack of real-time social contacts, serological data, and attitude/behavioural surveys was felt more frequently for all three kinds of outreach when compared to their relative reported missingness across all studies. However, Fisher’s exact test with Bonferroni correction (p > 0.05) did not indicate substantial differences in data missingness between studies with and without reported interactions with PHAs.

Our analysis of the public health impact of the modelling studies quantified by policy citations showed that 52% of the studies were cited 122 times in 103 unique policy documents from 17 countries, from the European region, and International Governmental Organizations in 9 different languages. Studies of the early- and mid-pandemic phases had more policy citations than studies on the later-pandemic phase (average number of policy citations per study 2.2, 2.0 and 1.3, respectively for early-, mid-, and later-pandemic phases, respectively, Figure S1). Studies aiming at anticipation of the pandemic trajectory and evaluating the impact of NPIs had, on average, more policy citations (2.8 and 2.9, respectively, Figure S1).

## Discussion

By evaluating COVID-19 outbreak analytics practices in four European countries and across different pandemic phases, we revealed that evolving and complex public health needs drove the scope of analytical efforts. Similar to previous public health emergencies,^30^ early modelling focused on understanding epidemic dynamics, such as transmission parameters and disease burden. Over time, attention shifted toward evaluating NPIs and, later on, vaccination strategies, as reported previously based on UK-based modelling efforts.^5^ Nonetheless, understanding SARS-CoV-2 transmission remained central throughout, as emerging viral variants and changes in affected population characteristics, such as waning and growing immunity, required an ongoing reassessment of key epidemiological parameters.

Evolving outbreak phases shape data use and requirements,^30,31^ with the long-term nature of the COVID-19 crisis revealing both advances and gaps in existing disease surveillance systems and data pipelines. We found that modellers’ data needs were met for ∼40% of the studies. Two-thirds of studies relied on surveillance data. Given the persistent need to reassess epidemiological parameters, case linelists, incidence rates, and genetic data remained fundamental requirements during all pandemic phases, as also corroborated by Jit and others, based on their reflections and experiences with COVID-19 pandemic modelling in Western European countries.^7^ Furthermore, fluctuating epidemic waves prolonged the need to (re-)assess optimal NPIs in a continuously changing epidemiological context. Among non-traditional data sources, mobility data proved critical, and we found that they were used in half of the studies. As PHA sought to balance health, social, and economic costs, non-traditional data sources such as real-time mobility,^e.g.32^ social contact data,^e.g.33^ and behavioural surveys became increasingly needed to anticipate complex interactions between the virus’ epidemiology and the behavioural response to interventions in a context of epidemic fatigue and misinformation spreading.

In some cases, real-time data collection and linkage were made possible by innovative coordination frameworks, built during the emergency and involving the integration of governmental bodies, academia, and the private sector. Examples include the REACT program to track the progress of England’s epidemic through home testing^34^ and the COG- UK (https://www.cogconsortium.uk/) and EMERGEN consortia for virus sequencing in the UK and France, respectively. Initiatives for collating, curating and standardising data from diverse sources, e.g. Global.health and ourworldindata.org made data readily available for modelling, limiting barriers and delays in data acquisition and processing on surveillance data. We found that for-profit organisations were the main source of mobility data and the second largest source of data overall by either selling or sharing the data directly with modellers under non-disclosure agreements (IATA, mobile phone companies, Google and Facebook) or making coarsely aggregated data openly available (e.g. COVID-19 community mobility reports by Google). Other relevant human behaviour data were collected by research consortia funded by the European Commission, e.g. CoMix, to collect real-time social mixing data in over 20 countries.^35^

Despite these efforts, our study showed that necessary data were often not available to modellers when needed, most notably real-time social contact and attitude/behavioural information, followed by serological data. In part this may be due to the time needed to secure funding and set up academic consortia (as these data were not routinely available before the pandemic), resulting in delayed availability of these data. Moreover, according to survey respondents, data were unavailable for sufficient locations under study and/or lacked essential stratification and standardisation. Kraemer and colleagues recently provided an overview of how advances and availability of artificial intelligence (AI) methods have the potential to more rapidly integrate disparate data sources, enabling rapid risk assessments and providing guidance for policymakers during health emergencies^36^. They are however dependent on the availability of robust data, which remains one of the main barriers for their deployment. During “peacetime”, enhanced data collection campaigns, as listed above, should be made protocol-ready and scalable to increase the geographical coverage, sustainable data collection and operational readiness for future pandemics. Similar to the First Few X cases (FFX) or the UNITY study protocols,^37^ standardised data collection protocols for non-traditional surveillance data could be formulated. This should be done by maintaining continuous collaboration between academic and public health sectors on the one hand and for-profit data providers on the other. Hence, restrictions, costs, and delays in data sharing during public health emergencies can be reduced.^38^

Existing participatory platforms similar to Influenzanet.info could be leveraged to deploy and scale up surveys rapidly. The Social Study (thesocialstudy.be), set up by Belgian Universities in collaboration with federal and local governments, aims to follow a representative group of community members and their sentiments and opinions on various topics by deploying surveys regularly in non-pandemic times. Such post-COVID-19 open-source data initiatives serve as valuable resources to further integrate behavioural and sentiment data in modelling frameworks and help mitigate threats arising from misinformation,^38^ and monitor risks.^39^

Analysing the modelling–PHA interface we found that in all four countries, modelling substantially informed COVID-19 pandemic decision-making. In particular, understanding the impact of NPIs was key to public health authorities’ needs. Indeed, studies with this aim were more represented among the studies that supported PHAs’ scientific understanding and were also among the ones, together with pandemic anticipation studies, that obtained more policy citations. We furthermore found an evolving integration of modelling into decision-making. Advisory committees were increasingly established during the mid- and later-pandemic phases, with their multi-disciplinarity growing to address growing complexities (e.g. the Belgian pandemic management committee, the French Covid-19 Scientific Advisory Board, and SPI-M-O in the United Kingdom) thus exchanges of requests and modelling frameworks used became more streamlined and transparent^7,24^. In the early phase of the COVID-19 crisis when epidemiological uncertainty was at its highest and public health decisions had to be made rapidly, informal networks of collaborations among modellers across European countries played a crucial role in facilitating the exchange of information, knowledge, and modelling approaches. These networks, often built through previous European Commission-funded consortia, enabled researchers to share real-time insights on epidemiological parameters, methodological approaches, and evolving data sources. Such pre-existing collaborations amplified the impact of formal structures and contributed to shaping early response strategies by leveraging shared expertise, thus reinforcing the importance of sustained investment in cross-border modeling networks for pandemic preparedness.

Building on this experience, national governments, including those in the EU/EEA, are investing in pandemic preparedness and resilience in the post-COVID era. They establish national and regional pandemic intelligence systems and networks of modellers and public health actors (e.g., European Centres of Disease Control’s respicast.ecdc.europa.eu and respicompass.ecdc.europa.eu, influcast.org, World Health Organisation’s epi-parameter community, the Dutch Institute of Public Health and the Environment’s modelling platform for policymakers, and the CDC’s Center for Forecasting and Outbreak Analytics). In a time where open source and open data are advocated, nearly half of the surveyed studies made code openly available. Delays in availability often stemmed from the need to prioritise rapid outputs for outbreak response over producing well-documented, usable code. Creating collaboration frameworks as described above and increasing incentives or academic credits for code- and data-sharing could facilitate faster and more flexible analytical framework and data sharing,^40^ which in turn can increase reproducibility and cross-country comparability.

A limitation, but simultaneously a strength of our work, is that data feeding into this study was provided by collaborators of modellers spanning four European countries. While this may not be representative for all of the EU/EEA, our policy analysis showed that the relevance and uptake of the described work reached a vast number of countries and regions. Secondly, non-traditional economic and proxy- or environmental data sources were not listed among the data sources used and missing, whilst these have reportedly been used to inform COVID-19 modelling work. This may relate to our standardised answers format, which lacks some of these mentionings. However, as we used a first-scoping survey to define our answer categories, we believe our responses are largely representative.

### Conclusion

Before and during the COVID-19 pandemic, substantial EU funding was directed to pandemic preparedness collaborations and research. By inventorying the contributions of a large Horizon2020 project, we outlined the pandemic response’s data and analytical needs while, at the same time, assessing science-policy interactions and uptake for pandemic response through such consortia and networks. Hence, we provide valuable insights that can support priority-setting for future surveillance investments and funding mechanisms for pandemic preparedness. A major challenge for outbreak analytics is the availability of data. In particular, integrating data on population characteristics, sentiment and human behaviour proved essential but also challenging components of mathematical models. Our approach can be further expanded to other geographical areas and emerging disease threats. Linking our approach with WHO’s Preparedness and Resilience for Emerging Threats (PRET) initiative, priority disease models based on modes of transmission can be defined and used to map further enhanced surveillance data needs, guide flexible analytical framework development, map actors and build collaborative networks.

## Data Availability

All data produced in the present study are available upon reasonable request to the authors

## Data sharing statement

The code to perform the analyses reported in the paper is available at https://github.com/chiara-poletto/Modelling-practices/.

## Acknowledgements

The work was supported by EU grant 874850 MOOD and is registered as MOOD publication 133. The contents of this publication are the sole responsibility of the authors and do not necessarily reflect the views of the European Commission. S.D. acknowledges support from the *Fonds National de la Recherche Scientifique* (F.R.S.-FNRS, Belgium; grant n°F.4515.22), from the Research Foundation — Flanders (*Fonds voor Wetenschappelijk Onderzoek — Vlaanderen*, FWO, Belgium; grant n°G098321N), and from the European Union Horizon 2020 projects MOOD (grant agreement n°874850) and LEAPS (grant agreement n°101094685). W.V.B. acknowledges funding from the Belgian Science Policy research grant BE-PIN (TD/231/BE-PIN) and is a member of the Outbreak Research Team (ORT) of the Institute of Tropical Medicine. This ORT is financially supported by the Department of Economy, Science and Innovation of the Flemish government. S.L.v.E. acknowledges funding from the MRC Centre for Global Infectious Disease Analysis (reference MR/X020258/1), funded by the UK Medical Research Council (MRC). This UK funded award is carried out in the frame of the Global Health EDCTP3 Joint Undertaking. V.C. acknowledges support from EU Horizon 2020 grant MOOD (H2020-874850), Horizon Europe grant ESCAPE (101095619), ANR grant DATAREDUX (ANR-19-CE46-0008-03). M.U.G.K. acknowledges funding from The Rockefeller Foundation (PC-2022-POP-005), Google.org, the Oxford Martin School Programmes in Pandemic Genomics & Digital Pandemic Preparedness, European Union’s Horizon Europe programme project E4Warning (#101086640), Wellcome Trust grants 303666/Z/23/Z, 226052/Z/22/Z & 228186/Z/23/Z, the United Kingdom Research and Innovation (#APP8583), the Medical Research Foundation (MRF-RG-ICCH-2022-100069), UK International Development (301542-403), the Bill & Melinda Gates Foundation (INV-063472) and Novo Nordisk Foundation (NNF24OC0094346). C.P. acknowledges funding from Cariparo Foundation through the program Starting Package. The funders had no role in the manuscript. The authors declare no conflicts of interest.

## Author’s contributions

E.v.K., C.P., W.V.B., and E.A. conceived the study. S.E. searched and extracted results from the Overton database, C.P. and E.v.K. performed all analyses and wrote a first draft of the manuscript, followed by inputs from W.V.B., E.A., and V.C. All co-authors filled out the survey and contributed to subsequent revised manuscripts, and provided discussion and comments. E.v.K. and C.P. are responsible for the content as guarantors.

