## Supplementary material for "Modelling practices, data provisioning, sharing and dissemination needs for pandemic decision-making: a European survey-based modellers’ perspective"

**Table of content**

### Scoping review of existing evidence

We conducted a systematic search in PubMed and MEDLINE between January 2020 and March 2024, using keywords related to SARS-CoV-2 (“COVID-19” OR “SARS-CoV-2” OR “2019-nCoV” OR “coronavirus”), mathematical modelling (model* AND “Mathematical” OR “Epidemiological” OR “Statistical” OR “Computational” OR “Bioinformatics”), and decision-making (“Policy” OR “Decision-making”) and no geographical limitations other than redistricting our search to English language. This search yielded 1,971 studies, of which 40 papers were shortlisted for full-text review, and 18 were in line with the scope of our study and one was identified through snowballing. These prior studies primarily concerned individual countries and/or qualitative accounts from predominantly earlier phases based on scoping reviews, interviews or published commentaries written by scientists involved in the COVID-19 response. Prior studies highlighted lessons learned related to relevant modelling frameworks and their data requirements, as well as data sharing and availability challenges, the importance of ensuring transparency, reproducibility and model validation for cross-country analyses, communicating model uncertainty and outcomes to policy-makers, and the role of structured collaboration between scientists and decision-makers.

This study provides a first systematic and quantitative evaluation of evolving data, analytical techniques, and science-policy interactions of COVID-19 modelling for pandemic decision-making across multiple EU/EEA countries and pandemic phases through EU-funded partnerships. The work presents an evaluation and analytical framework that combines all dimensions of outbreak analytics, which previous studies addressed in part, and can be adapted to other geographical contexts and future threat scenarios.

### Survey on European modelling practices, data provisioning, sharing and dissemination for pandemic decision-making

**General information**

**Email** …

**Partner** …

**Short title of the work** …

**Section 1 of 5: Scope of the modelling work**

**1.1 What was the objective of the modeling work?**

*Multiple answers allowed*

- anticipation – short term forecast
- anticipation – scenario analysis
- anticipation – risk of geographical spread
- anticipation – other
- evaluate impact npi – contact tracing
- evaluate impact npi – isolation/quarantine
- evaluate impact npi – lockdown
- evaluate impact npi – mask
- evaluate impact npi – school-based interventions
- evaluate impact npi – social distancing
- evaluate impact npi – screening
- evaluate impact npi – travel restrictions
- evaluate impact npi – relaxation/exit strategies
- evaluate impact npi – other
- Vaccination
- monitor cases
- understand the epidemic dynamics - emergence
- understand the epidemic dynamics - animal/human interface
- understand the epidemic dynamics - estimate transmission parameters
- understand the epidemic dynamics - determinants of geographical spread
- understand the epidemic dynamics - COVID-19 burden
- understand the epidemic dynamics - estimate true number of cases/under-reporting
- understand the epidemic dynamics - clinical aspects (e.g. risk factors, clinical manifestations)
- understand the epidemic dynamics - other
- social/behavioural response to COVID-19 pandemic
- early signal detection/monitoring
- improve data access/use
- covid impact on other public health issues
- Other …

**1.2 What was the geographical scope of the work?**

- Global
- Continent
- Country
- Within country
- City
- Not defined
- Other …

**1.3. Name the geographical territory** *(If not relevant, write “not relevant”)* …

**1.4. Could the study be easily adapted to another geographical territory?**

- Yes, everywhere worldwide
- Yes, in a similar socio-demographic contexts
- No

**1.5. Is the modeling work applicable to other diseases?**

- Yes
- No

**1.6. Pandemic period the study refers to** *(Month/year of beginning - month/year of end)* …

**Section 2 of 5: Data and Data Availability**

**2.1 What data was used in the study?**

*Multiple answers allowed*

- mobility data - pre-pandemic international mobility
- mobility data - real-time international mobility
- mobility data - pre-pandemic domestic mobility
- mobility data - real-time domestic mobility
- mobility data - mobility/presence data relative to the pre-pandemic period (e.g. COVID-19 Community Mobility Reports by Google)
- epidemiological data - cases’ line-list data / contact tracing / case investigation
- epidemiological data - incidence data
- epidemiological data - hospitalization
- epidemiological data - mortality
- epidemiological data - serological data
- epidemiological data - genomic data
- social structure (household structure, school catchments, workplace size and distribution)
- healthcare demographics (beds in ICU, number of healthcare workers, location of testing centers)
- population characteristics (age, gender, ethnicity, comorbidities)
- pre-pandemic social contacts
- real-time social contacts
- vaccination data
- intervention data (e.g. Oxford COVID-19 response tracker)
- media/microblogging
- animal behavior
- covid-19 epidemiological data in animal
- attitude/behavioral surveys
- no data
- Other …

**2.2 For each of the data used in the study, how was this data available to you?**

*Multiple answers allowed*

- data were purchased
- data were freely and openly available since before the pandemic
- data were shared with the collaboration by for-profit organizations
- data were made freely and openly available by for-profit organizations during the COVID-19 pandemic (e.g. COVID-19 Community Mobility Reports by Google)
- data were collated and made freely and openly available during the COVID-19 pandemic by initiatives of academics or nonprofit organizations (e.g. Johns Hopkins SARS-CoV-2 testing dashboard, Our World in Data)
- data were collected and shared in real time by public health authorities/decision makers
- data were collected by study collaborators
- not applicable
- Other …

**2.3 What data were missing that would have helped improve the study?**

*Multiple answers allowed*

- mobility data - pre-pandemic international mobility
- mobility data - real-time international mobility
- mobility data - pre-pandemic domestic mobility
- mobility data - real-time domestic mobility
- mobility data - mobility/presence data relative to the pre-pandemic period (e.g. COVID-19 Community Mobility Reports by Google)
- epidemiological data - cases’ line-list data / contact tracing / case investigation
- epidemiological data - incidence data
- epidemiological data - hospitalization
- epidemiological data - mortality
- epidemiological data - serological data
- epidemiological data - genetic sequences
- social structure (household structure, school catchments, workplace size and distribution)
- healthcare demographics (beds in ICU, number of healthcare workers, location of testing centers)
- population characteristics (age, gender, ethnicity, comorbidities)
- pre-pandemic social contacts
- real-time social contacts
- vaccination data
- intervention data (e.g. Oxford COVID-19 response tracker)
- media/microblogging
- animal behavior
- covid-19 epidemiological data in animal
- attitude/behavioral surveys
- no data were missing
- Other …

**2.4 Why were the data missing?**

*Multiple answers allowed*

- data existed and were available on purchase, but we lacked for financial resources to purchase them
- data existed but the process to obtain them was too lengthy or complicated
- data existed, but they were protected for privacy/ethical reasons
- as far as we know data did not exist
- not applicable
- Other …

**2.5 Which of the following limitations applied to the data used in the study?**

*Multiple answers allowed*

- data were not available for all geographical locations under study
- data became available too late
- data stopped being available too early
- data became available with a delay
- lack of standardization across regions
- lack of standardization over time
- lack of essential stratification (e.g. age, gender, vaccination status)
- lack of reference pre-pandemic data
- low data quality (e.g. extensive missing data, errors, biases)
- low spatial resolution - mobility data
- low spatial resolution - epidemiological data
- low spatial resolution - social structure/population characteristics
- low spatial resolution - healthcare demographics
- low spatial resolution - social contacts data
- low spatial resolution - vaccination data
- low spatial resolution - intervention data
- low spatial resolution - media/microblogging
- low spatial resolution - animal data
- low spatial resolution - attitude/behavior surveys
- low temporal resolution - mobility data
- low temporal resolution - epidemiological data
- low temporal resolution - social structure/population characteristics
- low temporal resolution - healthcare demographics
- low temporal resolution - social contacts data
- low temporal resolution - vaccination data
- low temporal resolution - intervention data
- low temporal resolution - media/microblogging data
- low temporal resolution - animal data
- low temporal resolution - attitude/behavior surveys
- no limitations
- Other …

**Section 3 of 5: Modelling Approaches and Availability**

**3.1 Modeling approach used**

*Multiple answers allowed*

- compartmental modeling
- statistical modeling
- descriptive statistics
- parameter fitting
- agent-based modeling
- metapopulation modeling
- data visualization/mapping
- network analysis
- phylogenetics/phylodynamics
- Bioinformatic
- web scraping/data mining
- machine learning
- Other …

**3.2 Did you use a code openly available?**

- Yes
- No

**3.3 Did you make the code openly available?**

- Yes
- No

**3.4 If you made the code openly available add here the link to the code repository** …

**Section 4 of 5: Use and role of modelling work**

**4.1 Was the study prompted by discussion with Public health authorities/decision makers?**

- Yes
- No

**4.2 Did the study support official recommendations by public-health authorities/decision makers?**

- Yes
- No

**4.3 Did the presented modeling influence the scientific understanding for situational**

**awareness of public-health authorities/decision-makers?**

- Yes
- No

**Section 5 of 5: Interaction and communication**

**5.1 Describe the interaction with public health authorities/decision makers in the context of the study**

*Mark only the closed interaction. For instance, if the interaction occurred through discussion at internal meeting and public health authorities were also involved in the collaboration mark "direct collaboration"*

- direct collaboration
- discussion at internal meeting
- interaction within advisory committees
- no interaction
- Other …

**5.2 Means by which the study was communicated to public health authorities/decision makers?**

*Mark only the more direct means of communication. For instance, if the results were directly share by email and the work was presented at workshop/conference where public health authorities were present mark "results shared by email"*

- direct collaboration
- discussion at internal meeting
- presentation within advisory committees
- results shared by email
- presentation at workshops/conferences
- scientific report/publication, but no direct interaction
- Other …

**5.3 Reference type**

*In case the same work was shared by different means, e.g. report and scientific publication, indicate all of them*

- internal report
- published report
- Pre-print
- scientific publication
- media dissemination
- Other …

**5.4 Reference url** …

**5.5 Reference title** …

**5.6 Reference date** …

**5.7 Size of the collaboration**

- the study involves less than five institutions
- the study involves more than five institutions

### Additional results

#### Policy impact of modelling studies

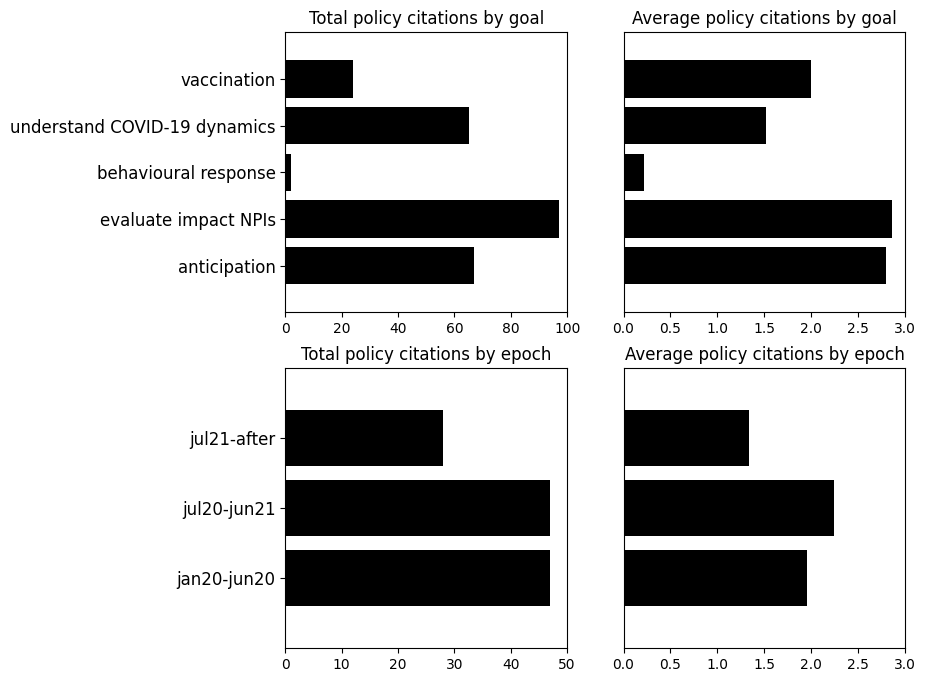

**Figure S1. Public health impact of modelling studies - stratified analysis. *Top left,*** *Number of policy citations of surveyed studies by goal of the study.* ***Top right,*** *Average number of policy citations of surveyed studies by goal of the study.* ***Bottom left,*** *Number of policy citations of surveyed studies by study epoch.* ***Bottom right,*** *Average number of policy citations of surveyed studies by pandemic period.*
